# FREQUENCY OF INFECTION AFTER CEREBROSPINAL FLUID SHUNTING PROCEDURES

**DOI:** 10.1101/2020.04.01.20050096

**Authors:** Kashif Ramooz, Eesha Yaqoob, Nadeem Akhtar, Fraz Mehmood, Saad Javed

## Abstract

Hydrocephalus is routinely treated by surgical procedures. Cerebrospinal fluid shunt placement is a critical therapeutic intervention for hydrocephalus.CSF shunting has multiple complications among which infection is very common. The major cause of morbidity and mortality in patients with CSF shunts is theinfection of the central nervous system (CNS).It can lead to prolonged hospital stay, increase the number of operative procedures 03 times more than then none infected cases and has twice the fatality rate. Study of such type of complication will help the patients to improve their health and also improve our sterilization techniques and reduce burden of hospital and patients expenditures. The objective of the study was to determine the frequency of infection after cerebrospinal fluid shunting procedures.Case series study was used as study design.Study was conducted from 10-2010 to 10-06-2011.One hundred and forty four patients with both genders of all age groups undergoing cerebrospinal fluid shunting, meeting inclusion and exclusion criteria, were selected for the present study after informed consent of patient or guardian and approval by the hospital ethical committee. Follow up was ensured by taking the telephonic contact and address of patient.Total no of patients were 144 among which, 89 were males and 55 were females. Age distribution was from 01 month to 75 years with the mean age of 15.280 and standard deviation was ± 20.450. Post-operative infection was present in 20(13.9%) patients.

**Author’s approval:** All the authors have seen the manuscript and approved it.

**Declaration of interest:** **None**

**Conflict/Competing of Interest:** **None**.

**Disclosure of Funding:** **None**.

**Ethical Approval:** **Attached**

## INTRODUCTION

The eradication of infection in patients with colonized shunts has always been a great challenge to the treating surgeon. Various methods have been tried with variable success. Meticulous aseptic measures, preoperatively and pre-operatively are essential in preventing colonization [1].Shunt infections occur generally within two months of shunt insertion and most common organism is staphylococcus epidermis.Despite the high incidence of infection, optimal management is still debatable, and research on prevention has been hampered by single-institution series and small numbers [2].CSF shunting has various complications among which shunt infections is the commonest [3].

As data is not available, it is not possible to make early diagnosis of shunt infection in patients with nonspecific clinical features. Among common predisposing factors for shunt infection are infants with <6months of age, immunologic deficiency and with their residential bacterial flora have higher incidence [4].

During early hydrocephalus studies, researchers investigated hydrocephalus by ventricular and lumbar punctures. By these methods some researchers injected supra vital dye into ventricles and then performed lumbar punctures. If spinal tap was done and dye detected, the hydrocephalus was called communicating, if not then hydrocephalus was called non-communicating or obstructive [5].

Treatment options for hydrocephalus are expensive for poor people. Due to limited facilities in remote areas, patient^’^s access to neurosurgical setup is difficult. For these reasons, treatment of hydrocephalus is delayed and frequency of hydrocephalus is higher in developing countries [6].

Mostly, hydrocephalus is developed due to disturbance in the absorption and drainage in the blood stream. This can happen due to multiple reasons as hemorrhagic stroke in adult patients cause hydrocephalus due to impaired absorption of CSF. Also meningitis causes resistance in absorption and drainage of CSF resulting in hydrocephalus. Rarely hereditary elements will cause hydrocephalus [7].

Approximately 10% of CSF shunt procedures are associated with infection and require removal followed by insertion of new shunt after treatment with antibiotics.Presenting features to diagnose shunt infection are fever, altered sensorium, seizures and abdominal fullness. In the adults, common features of shunt infection are fever and disturbed sensorium while in children disturbed sensorium and abdominal fullness are very common [8].

In this condition, pathologic enlargement of the cerebral ventricles occur secondary to increased intracranial pressure caused by a disturbance in the secretion, flow or absorption of cerebrospinal fluid (CSF).CSF is produced by the choroid plexus of the two lateral, third and fourth ventricles though some may be formed on the surface of the brain and spinal cord [9].Colonized shunts cannot function well mechanically [10].

Other methods used to reduce the CSF production have been irradiation of the choroid plexus and pharmacological therapy with Isosorbide or Acetazolamide [11].

So, it is evident that hydrocephalus is a syndrome in which there is disturbance in the dynamics of cerebrospinal fluid due to different types of diseases. There were different potential sites at which CSF flow obstruction occurs and they also thought that all hydrocephalus are obstructive in nature [12].Hydrocephalus is a condition in which excess fluid accumulates in the brain [13].

Antibiotic-impregnated catheter shunt systems, in particular, appear to reduce infection risk in some, but not all, reported series. Unfortunately, none of these studies were performed in a prospective, double-blinded, randomized controlled fashion [14].Infection of the central nervous system (CNS) is a major cause of morbidity and mortality in patients with CSF shunts [15].

During the eighteenth and nineteenth centuries, diets and dehydration cures were suggested [16]. Long-term follow-up of shunted children is necessary to evaluate the real incidence of shunt Infection and the functional outcome after shunt Infection [17].

## OBJECTIVEOF STUDY

- To determine the frequency of infection after cerebrospinal fluid shunting procedures.

## MATERIALS AND METHODS

### STUDY DESIGN

Case series study.

### DURATION OF STUDY

Eight months (16^th^ October 2010 to 16 June 2011)

### SAMPLE SIZE

Total 144 patients were taken for this study among which 89 were males and 55 were females. All of which operated for CSF shunts.

### SAMPLING TECHNIQUE

Non-probability consecutive sampling

### SAMPLE COLLECTION

#### Inclusion criteria

1. All cases with communicating and non-communicating hydrocephalus.
2. All age groups with both genders.
3. Cerebrospinal fluid shunting under same type of anesthesia and antibiotic cover.

#### Exclusion criteria

1. Diagnosed case of active meningitis.
2. History of previous cerebrospinal fluid shunting.
3. Patients having co morbid conditions like diabetes mellitus, end stage renal disease, ischemic heart diseases, malnutrition and cirrhosis of liver.

### DATA COLLECTIONPROCEDURE

Patients with both genders of all age groups underwent cerebrospinal fluid shunting, meeting inclusion and exclusion criteria have been selected for the present study after informed consent of patient or guardian and approval by the hospital ethical committee. Operation has been done by consultant neurosurgeon. Pre and post-operative antibiotic prophylaxis such as Oxidil (Sammi), Vanccare (Citicare Laboratories Pvt. Ltd.) and Flagyl (Pfizer) has been given according to CSF shunt (Medtronic USA VP shunt medium pressure) protocol. Every patient has been examined clinically for infection by the trainee Researcher on 3^rd^, 7^th^and 15^th^ postoperative day. Clinically suspected cases have been confirmed by blood C-reactive protein, cerebrospinal fluid analysis and complete blood profile with ESR. Patients with no complications have been discharged on 3^rd^post-operative day and kept on follow up till 15^th^ postoperative day to consider complication free patient. Follow up has been ensured by taking the telephonic contact and address of patient.

### DATA ANALYSISPROCEDURE

Data has been analyzed using software named “Statistical Package for Social Sciences” version 11. Frequency and percentages have been calculated for qualitative variables like infection, gender and type of hydrocephalus. Mean and standard deviation has been calculated for quantitative variables i.e. age.

## RESULTS

There were total 144 patients in which CSF shunting was done for hydrocephalus from 16-10-2010 to 16-06-2011.

### Diagnosis of patients

Among 144 numbers of patients, maximum number of cases operated for non-communicating and communicating hydrocephalus was congenital hydrocephalus and minimum number of cases wascerbellopontine angle tumors.

**Table 1:**
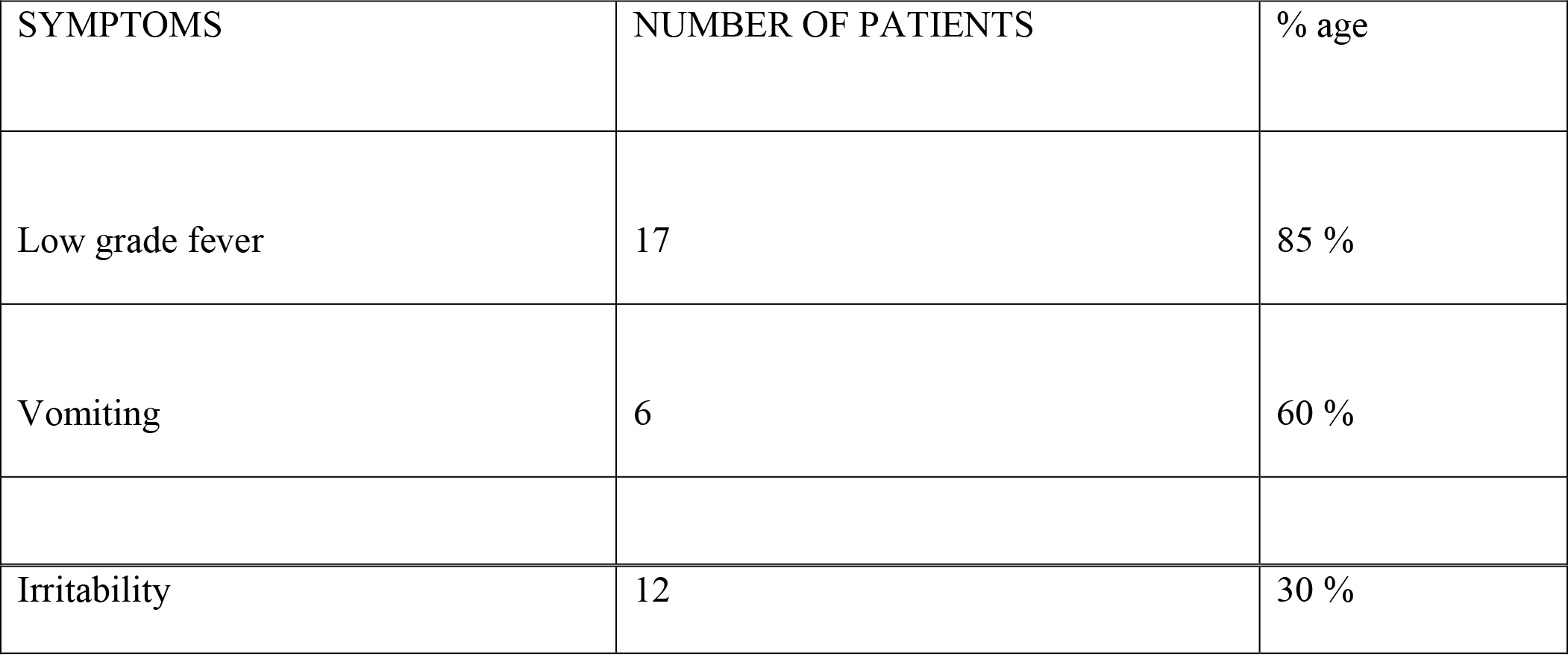
Common symptoms of Post-operative Infection (n=144) Values in table 1 showed that 85% of patients were low grade fever while 60% of patients faced the problem of vomiting and 30% of patients had the symptom of irritability. Table no 1showed that 85% of patients were low grade fever while 60% of patients faced the problem of vomiting and 30% of patients had the symptom of irritability.

| SYMPTOMS | NUMBER OF PATIENTS | % age |
| --- | --- | --- |
| Low grade fever | 17 | 85 % |
| Vomiting | 6 | 60 % |
| Irritability | 12 | 30 % |

**Figure 1.**
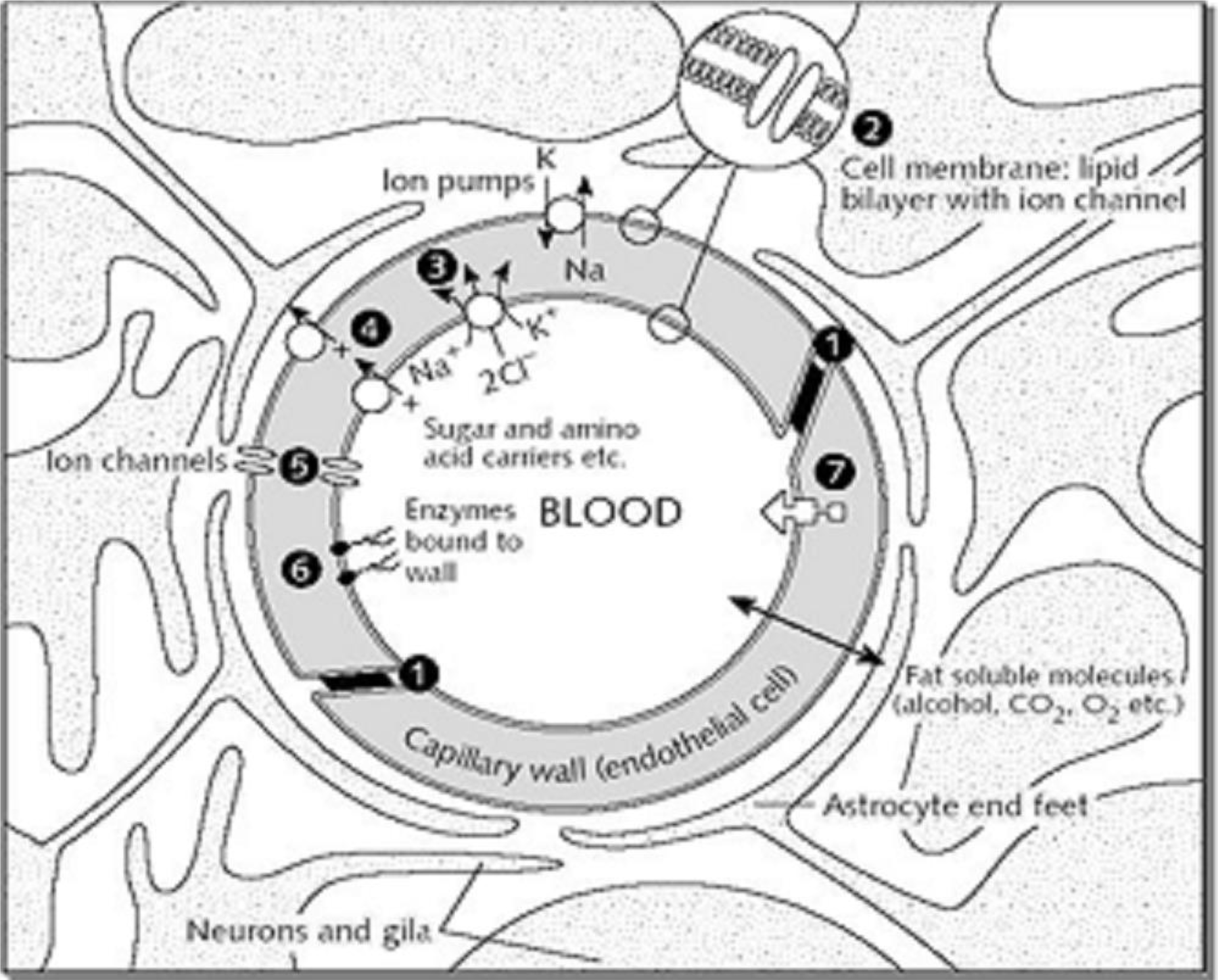
Diagram of a cerebral capillary enclosed in astrocyte end-feet. Characteristics of the blood-brain barrier are indicated: (1) tight junctions that seal the pathway between the capillary (endothelial) cells; (2) the lipid nature of the cell membranes of the capillary wall which makes it a barrier to water-soluble molecules; (3), (4), and (5) represent some of the carriers and ion channels; (6) the ‘enzymatic barrier’ that removes molecules from the blood; (7) the efflux pumps which extrude fat-soluble molecules that have crossed into the cells.

**Figure 2.**
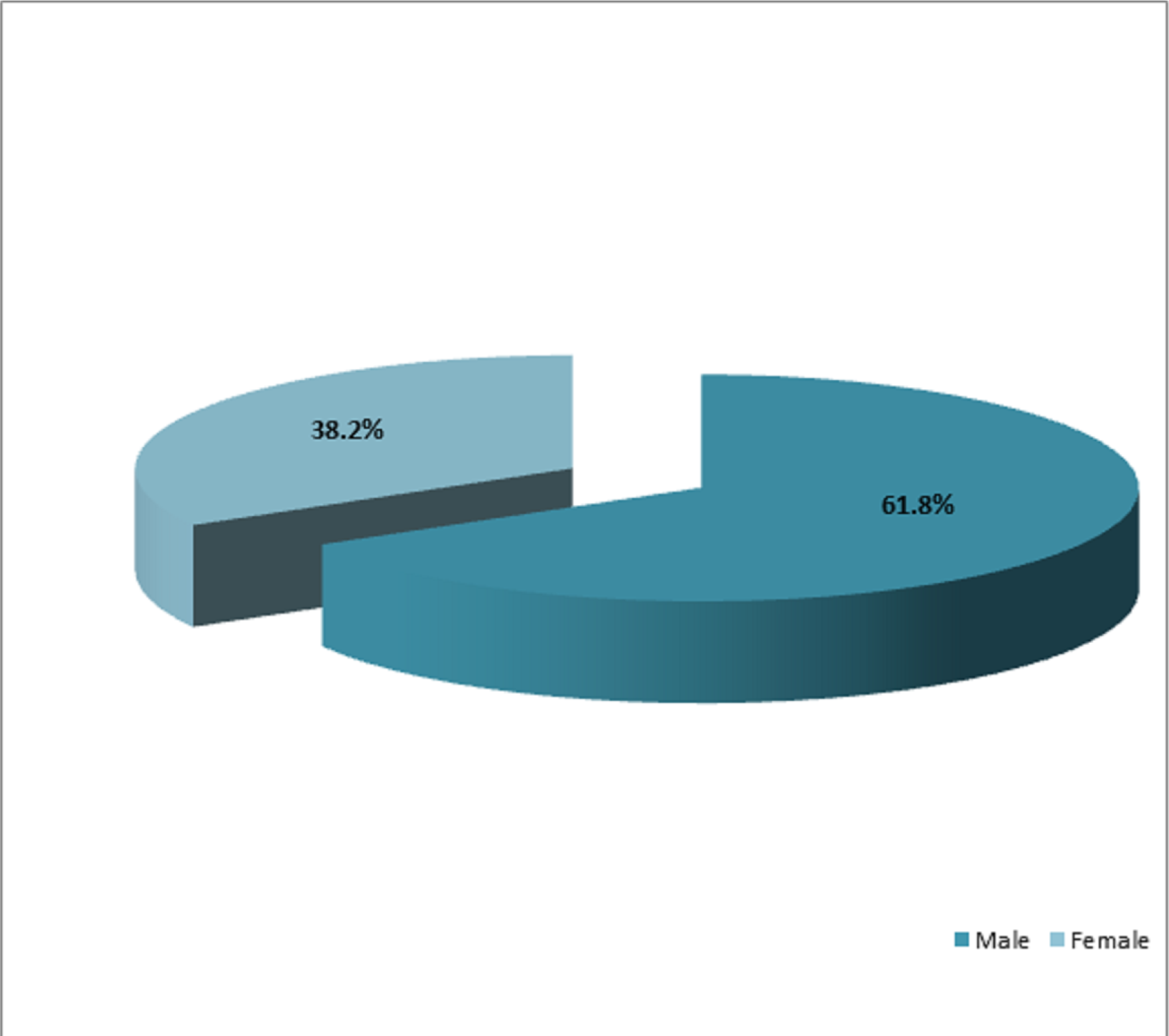
Out of these 144 patients, there were 89 (61.8%) males and 55 (38.2%) females with the ratio of 1: 0.6.

**Figure 3.**
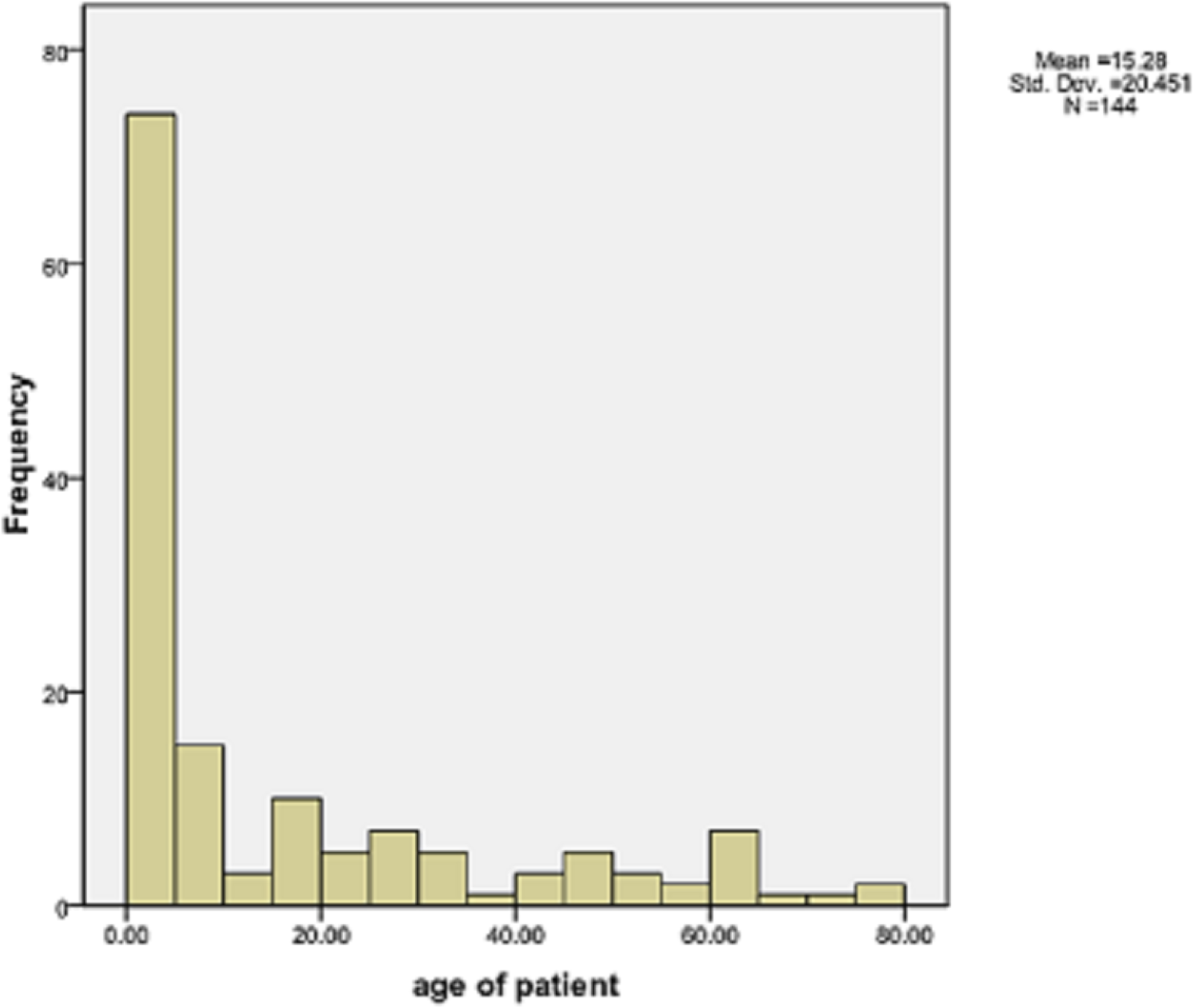
Patients were between the ages of 01 month to 75 years with the mean age of 15.280 and standard deviation was ± 20.450. Maximum age was 75 years and minimum was 01 month but no patient was under 1 month of age at operation. Highest number of patients was between the ages of 01 month to 07 years. Lowest figure was between the ages of 16 to 45 years.

### Clinically confirmed infection in post-operative periods

Among 144 patients, 20 (13.9%) had post-operative infection while 124(86.1%) had no evidence of infection. The infected patients had the symptoms of low grade fever (85%), irritability (60%) and vomiting (30%).

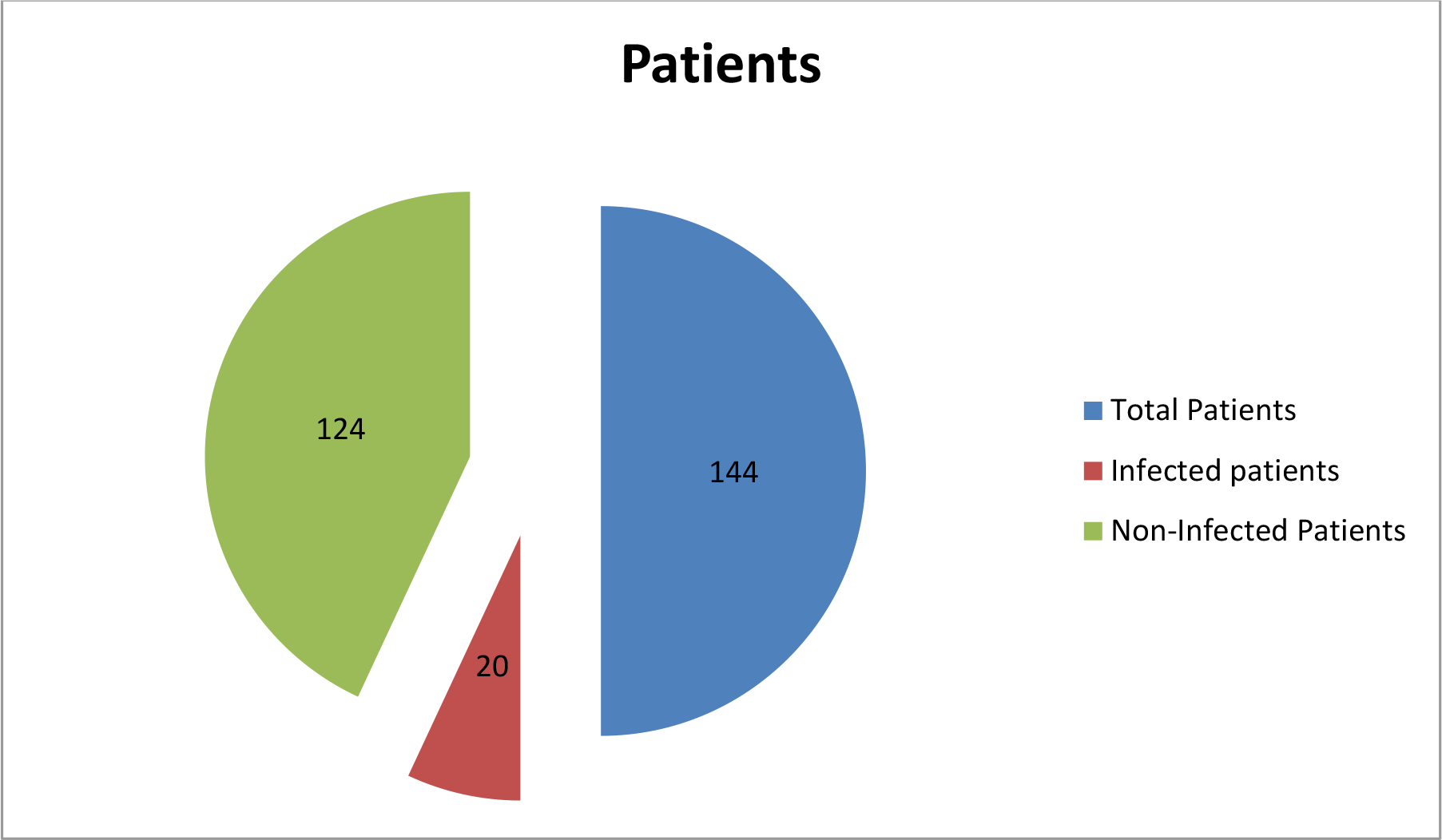

#### Laboratory findings

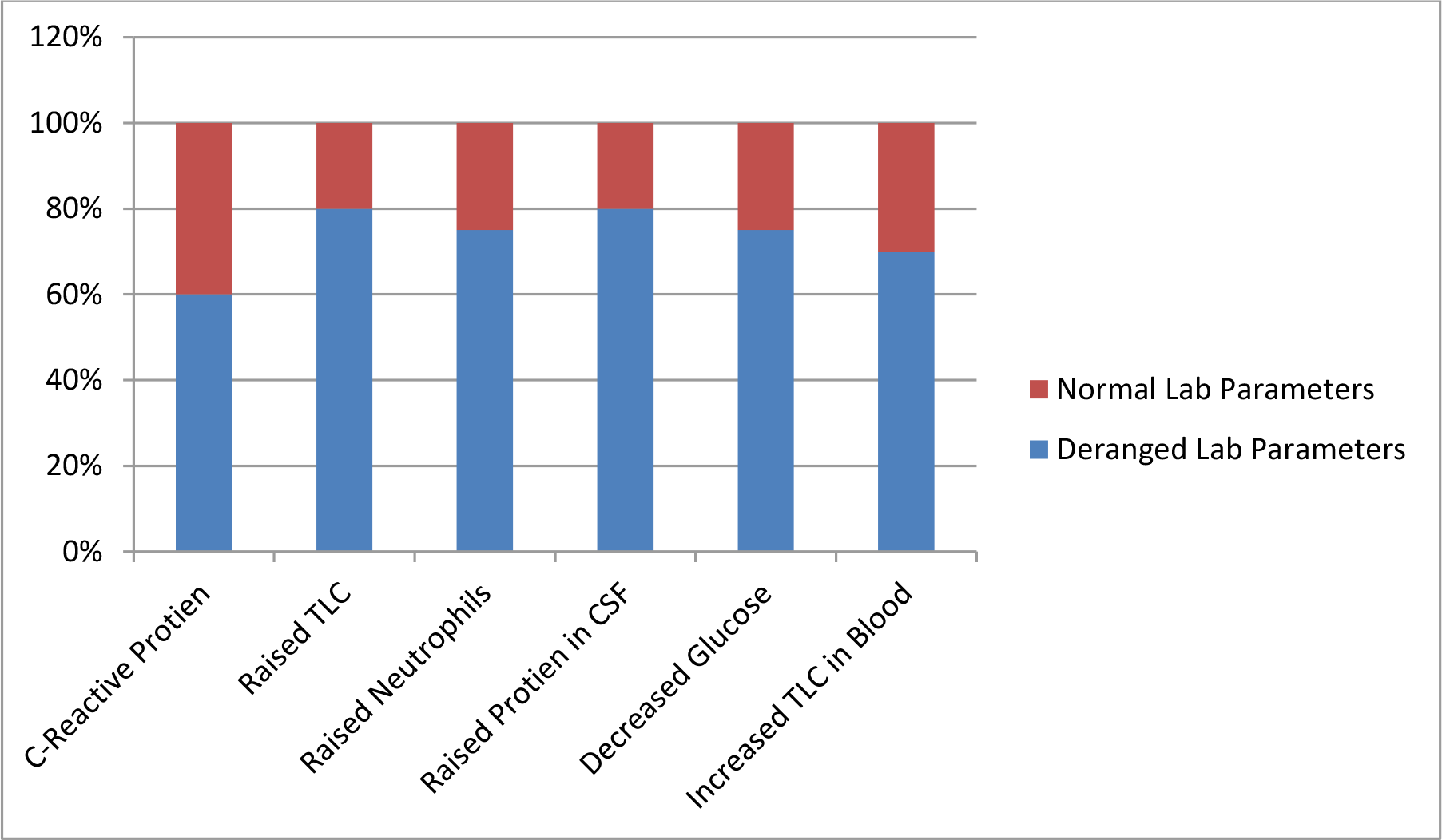

Post-operative 60% infected patients had C-reactive protein of more than 10mg/dl while some infected and all non-infected patients had value of 10 or less than 10mg/dl. Among these, 80% infected patients had raised TLC, 75% had raised neutrophils and 80% had raised protein in CSF. 75% had decreased glucose in CSF. 70% infected cases also had increased TLC in blood.

## DISCUSSION

The purpose of study was to determine the frequency of infection after cerebrospinal fluid shunting procedures and to improve our sterilization techniques, patients’ health and to reduce the burden of hospital, patient^’^s expenses and hospital stay.

In this study total number of cases were144 who were treated for hydrocephalus with CSF shunts. Patients were between the ages of 01 month to 75 years with the mean age of 15.4197± 20.820 and minimum age was 01 month and maximum was 75 years. Highest number of patients was between the ages of 01 month to 07 years. Lowest figure was between the ages of 16 to 45 years. Maximum number of patients had congenital hydrocephalus (26.7%), then patients with blocked shunts (20%). Among 144 patients, 20 (13.9%) had post-operative infection while 124(86.1%) had no evidence of infection. The infected patients had the symptoms of low grade fever, irritability and vomiting and maximum had showed high value of C-reactive protein considering the normal rang 0-10. All those infected cases were confirmed by laboratory investigations of CSF and blood. These infected patients had raised TLC, neutrophils and protein in CSF while they had decreased glucose in CSF. Also there was raised TLC and ESR in blood.

Two hundred and twenty six cerebrospinal fluid (CSF) shunt related procedures performed. During the study period, nine shunt infections resulted from 226 shunt procedures, giving an overall infection rate of 3.98%. Of the nine patients who got infected, there were seven males and two females. Six of the males and one female were below 15 years of age. In this study, infection rate was very low as compared to our study.

## RECOMMENDATIONS

This study helps us to improve patients’ health and to reduce the burden of hospital, patient’s expenses and hospital stay by improving our sterilization techniques, prophylactic antibiotics and proper follow up.

## Data Availability

All the data is available with me and can be produced whenever required by the editor

## DECLARATION

- Ethics approval and consent to participate: Taken from the department and chair
- Consent for publication: All authors consented for publication
- Availability of data and material: available
- Competing interests: none
- Funding: none

## Authors’ contributions

Substantial contributions to the conception or design of the work; or the acquisition, analysis, or interpretation of data for the work: Dr Kashif Ramooz, Dr Saad Javed, Eesha Yaqoob

Drafting the work or revising it critically for important intellectual content: Dr Saad Javed Final approval of the version to be published; Eesha Yaqoob

Agreement to be accountable for all aspects of the work in ensuring that questions related to the accuracy or integrity of any part of the work are appropriately investigated and resolved: Dr Saad Javed, Eesha Yaqoob

## Acknowledgements

For the Rawalpindi medical university which gave us the platform to work and research

## Data availability statement

All the data is available with me and can be produced whenever required by the editor

